# Modelling, Extrapolation and Simulation approaches for rare diseases - Review of two case-studies in the INVENTS project

**DOI:** 10.1101/2025.09.17.25335979

**Authors:** Emmanuelle Comets, Moreno Ursino

**Affiliations:** Université Paris Cité, Université Sorbonne Paris Nord, Inserm, IAME, F-75018 Paris, France; Univ Rennes, Inserm, EHESP, Irset UMR_S 1085, F-35000 Rennes, France; Inserm, Université Paris Cité, Inria, HeKA, F-75015 Paris, France; Novartis, Basel, Switzerland; Pharmetheus, Paris, France; Roche, Basel, Switzerland; Uppsala University, Uppsala, Sweden; Inserm, Paris, France; Roche, Paris, France

**Keywords:** Longitudinal data, secukinumab, tocilizumab, literature review, modelling, simulation extrapolation

## Abstract

**Objective:** The objective of this work was to review the modelling and simulation methods used in two selected case-studies from the Invents project, with a focus on longitudinal data modelling, simulation methods and extrapolation approaches (across populations, across indications or across similar drugs) applied during the later phases of drug development.

**Methods:** We included publications based on relevance to the case-studies involved in Invents, extending the search to articles reporting results of modelling, simulation, or extrapolation approaches for the two drugs secukinumab and tocilizumab through keyword-based literature searches.

**Results:** When outcomes were measured repeatedly during the study, the focus of the analyses was usually along the lines of improvement at a given time-point, with other time-points included in secondary analyses. When PK (pharmacokinetic) or PK/PD (pharmacodynamic) models were developed, they were generally simple, with compartmental models for PK and direct or indirect response models for PD. Clinical questions addressed in the studies selected in this review were mainly centered on efficacy (57%, including exposure-response relationships and efficacy concerning patient-reported outcomes) and safety (34%). Besides the few studies investigating PK, other clinical questions focused on finding predictors of response, either biomarkers related to changes in the outcome under study, molecular characteristics of certain groups of patients, or prognostic factors at baseline. Finally, a couple of studies were more methodological. We found almost no paediatric study, despite modelling and simulation analyses being put forward in the Paediatric Investigation Plan, which could be due to the time needed to perform these studies, lack of interest in obtaining paediatric approval, waivers or failure to publish.

**Conclusion:** Despite the clear focus on longitudinal modelling and simulation in our search, we found very few examples of published PK or PK/PD models for the two case-studies considered, and almost no extrapolation approaches for paediatric or disease applications.

## 1 Introduction

Workpackage 1 in the INVENTS project (European Union’s Horizon Europe Research and Innovation programme, grant agreement 101136365) focuses on the analysis of longitudinal data, with a global aim of improving the robustness of extrapolation, modelling and simulation methods, and evaluation assessment criteria. Task 1.1 was defined to determine commonly used methods, approaches, evaluation procedures for longitudinal data modelling for the evaluation of the associated uncertainty and its propagation, and to identify the methodological gaps in these approaches and their assessment, focusing on two WP7 use cases. The methods and models could then be used to define scenarios for the simulation of virtual populations, propose clinical questions to address the definition of estimands, or test new methodological developments, while the gaps identified either in the methods or in their evaluation would serve as a benchmark to propose new approaches and new criteria in the other tasks of WP 1.1.

Our points of interest were modelling and simulation approaches (including type of model, estimation approaches, handling of missing data, relationship to the final clinical endpoints via dose/exposure-response models, etc.), the trial and study designs the models were developed on (population, dose-escalation, dose range, etc.), simulation procedures used (endpoints, sample size, biomarkers, handling of uncertainty, designs) and extrapolation approaches (allometric and maturational scaling, multiscale models, disease similarity). Clinical questions could involve improving the precision of the estimate of treatment efficacy, supporting extrapolation to a new and rare indication with limited data collected; revisit an existing and successful paediatric extrapolation in order to increase efficiency; identifying target endpoints earlier; extrapolate missing data (e.g. missing endpoints); quantifying the strength of evidence and assessing the additional information needed in order to validate the (model-based) evidence analysis.

The objective of this work was to review the modelling and simulation methods used in the two case-studies, with a focus on longitudinal data modelling, simulation methods and extrapolation approaches (across populations, across indications or across similar drugs) applied during the later phases of drug development.

We performed a case-study based search, first selecting material through the owners of the case-study and then expanding through keyword searches of the publication database PubMed and the references in the papers read in the survey. An extraction template was defined, and the papers collected in the survey were read by one reader each. This paper presents the results of the quantitative review, summarising the information obtained from the list of articles read.

## 2 Methods

### Scope

This review focuses on modelling, simulation, or extrapolation approaches published for the case-studies included in the INVENTS project. For tocilizumab, we considered publications in rheumatoid arthritis, the rare diseases giant cell arteritis and systemic sclerosis, and juvenile idiopathic arthritis as the paediatric form of the disease. For secukinumab, we considered publications in psoriasis and arthritis.

#### 2.1 Research questions

We aimed at answering the following research questions:

- What were the clinical questions of interest ?
- What type of data was available and at what stage ?
  – study design (number of patients, number and timing of samples, patient stratification)
  – treatment (single versus combination therapies, SoC,…)
  – prior knowledge
- What type of analysis was performed ?
  – standard statistical analyses (endpoint comparison, target achievement)
  – modelling and simulation studies
  – question of interest, estimands
- What types of longitudinal models have been developed to describe the data?
- What covariates have been used ?
- What types of extrapolations were made? E.g. between populations with
  – differences in patient characteristics (age, ethnicity (?), …)
  – differences in disease characteristics (disease severity, juvenile versus rheumatoid arthritis, progression rate,…)
- What simulation studies have been performed ?
  – what were the objectives ?
  – what models have been used (including covariates) ?

#### 2.2 Literature search

For all the searches described below, articles available from literature databases were stored in the Sharepoint for the WP 1 workpackage, as well as publicly available reports.

### Search Strategy

An initial list of articles was established by the proponents of each case-study to form the core basis of the articles searched. The full-text of the papers and reports in this first list were obtained and read by one reviewer.

We included articles and records of RCT published up to November 2024 on ClinicalTrials.gov related to the phase II and III clinical trials included in the data package in Invents for the two case-studies:

- Secukinumab
  – JIA: NCT03031782
  – PsA: NCT01392326, NCT01752634, NCT01989468, NCT02294227, NCT02404350
  – Psoriasis: NCT01365455, NCT01358578, NCT01555125, NCT01636687, NCT02471144, NCT03668613
- Tocilizumab
  – 4 phase III RCT in RA: NCT00106548, NCT00106574, NCT00106535, NCT00106522

For both drugs, we also considered publications cited in:

- regulatory reports (FDA and EMA reports, paediatric investigation plans)
- internal reports including modelling, simulation or extrapolation approaches, through a list established by each case-study owner

We excluded articles where the abstract clearly stated that no longitudinal modelling was present.

This initial search was then extended in September 2024 to a keyword search (see below), to access more papers on modelling and simulation. We then performed a final search in February 2025 targeting specifically pharmacokinetic and pharmacodynamic models in the pathologies considered in the case-studies.

### Study objectives

We included studies reporting either one of:

- Longitudinal data analyses (learn), which must contain one or more of the following:
  – modelling
  – simulation
  – extrapolation (paediatric, other disease,..)
- Pivotal studies (confirm), which must contain:
  – clinical questions
  – statistical analyses

All types of patients could be included (men, women, children, elderly, etc.).

#### 2.3 Template extraction

A data extraction form was developed in order to extract information about:

- Presence of modelling, simulation and/or extrapolation approaches
- Clinical questions addressed or asked, eg
  – improve the precision of the estimate of treatment efficacy
  – support extrapolation to a new and rare indication with limited data collected
  – revisit an existing and successful paediatric extrapolation in order to increase efficiency
  – identify target endpoints earlier
  – extrapolate missing data (e.g. missing endpoints)
  – quantify the strength of evidence and assess the additional information needed in order to validate the (model-based) evidence analysis
- Data
  – population (age range, special population)
  – main disease for which the drug is considered
  – inclusion criteria
  – exclusion criteria
  – covariates recorded (including time-varying covariates, comorbidites, concomitant treatments)
- Longitudinal endpoints
  – relationship to the final clinical endpoints
  – handling of missing data
- Modelling methods
  – model nature and scope
  – estimation methods
  – model building and covariate selection procedures
  – evaluation
- Extrapolation approaches
  – allometric and maturational scaling
  – disease similarity
  – multiscale models
- Trial and study designs
  – dose-escalation, dose range
  – study design, number of subjects, administration schedule
  – data collected
- Simulation procedures
  – biomarkers, endpoints
  – sample size
  – handling of uncertainty
  – design

Data extraction forms were designed to capture the information listed above and were used for the selected articles. All papers in the final database were read by one reader each, then sanitised and checked by EC during the data analysis step.

#### 2.4 Data analysis

The data analysis was descriptive including:

- A flow chart describing the list of publications found and selected for the quantitative analysis.
- Tables and figures with the characteristics of the included publications
- Tables and figures describing modelling approaches, simulation approaches, and extrapolation methods

The analysis was performed in R, using packages xtable, treemap, and ggplot2.

## 3 Results

### 3.1 Article selection

Figure 1 shows the selection and reading process. The first round included 55 papers, after which two additional rounds of keywords searches were added: 51 papers using keyword searches focusing on model and simulation, and 58 papers using keyword searches focusing on the main pathologies for each drug. Details are given in Supplementary table 2. For tocilizumab, a wider search (Search 0 in the supplementary table) triggered over 300 articles, most of which were irrelevant, so only the more specific searches were combined to be included in the final list.

**Figure 1:**
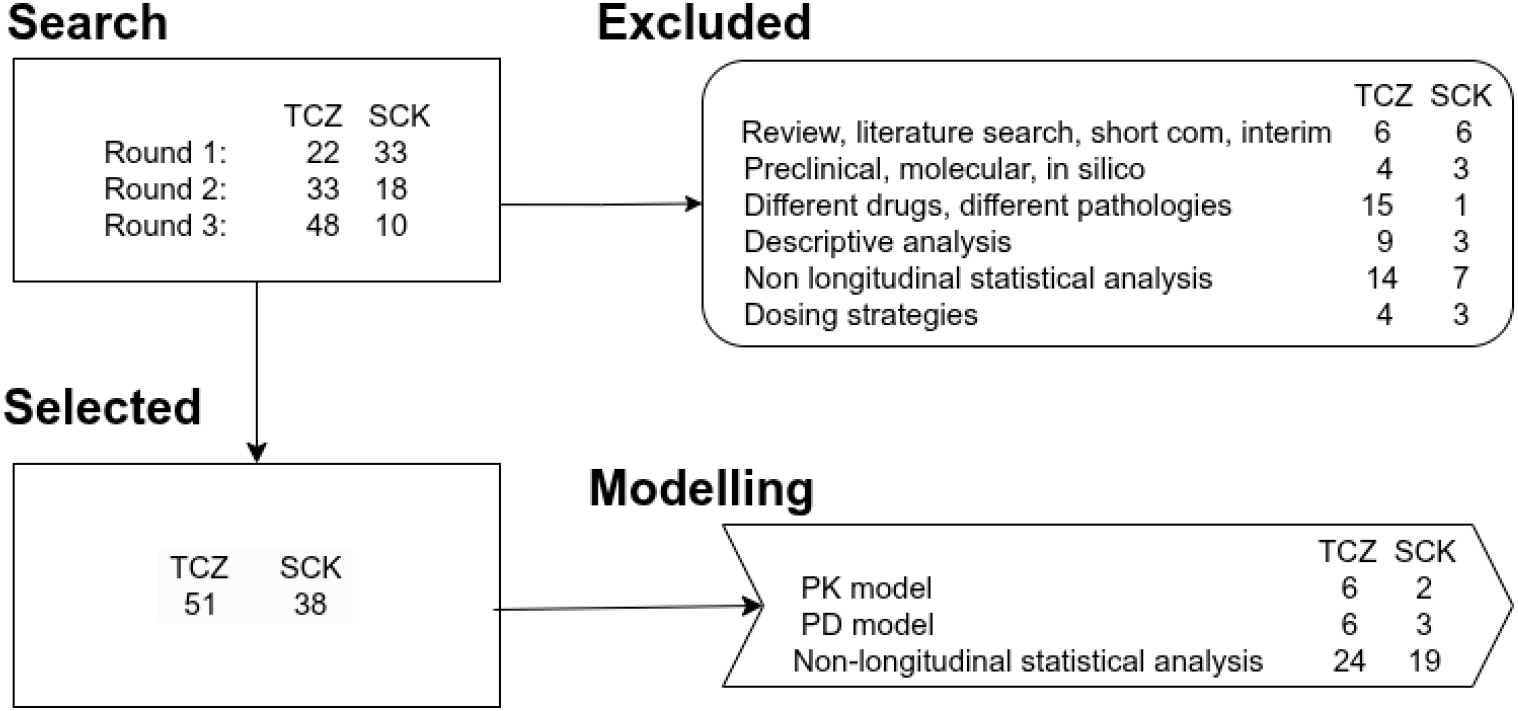
Selection process and articles read. In the modelling section, multiple choices were possible.

From the list of papers resulting from the searches, we excluded 76 papers, as shown in the flowchart. We excluded letters, short communications, and reviews, as well as preclinical or molecular modelling. Some papers were excluded because they concerned different drugs or pathologies, or compared treatment strategies with different drugs. We also excluded papers which were clearly descriptive in nature or presented only non-longitudinal statistical analyses. The final database included 89 papers, 51 for tocilizumab and 38 for secukinumab. Of these, only a few reported PK and/or PD modelling.

#### Article type

Figure 2 provides a histogram of the year the articles were published. The colours represent the articles with an author from the company developing the drug. Tocilizumab was first approved for rheumatoid arthritis in 2009 (EU) and 2010 (USA), and extended to sJIA in 2011, to pJIA in 2013 in both regions, as well as to giant cell arthritis in the United State in 2017. while secukinumab was first approved in plaque psoriasis in 2015 in the United States and in the EU. It was then approved for ankylosing Spondylitis and psoriatic arthritis in 2016 the next year. For secukinumab, most of the papers published had at least one clearly identified co-author in Novartis (34 out of 38 papers), while only 21 out of 52 for tocilizumab included a Roche author, with a larger proportion at the beginning of development when data was limited to in-house studies, and more papers published without ties to Roche in later years.

**Figure 2:**
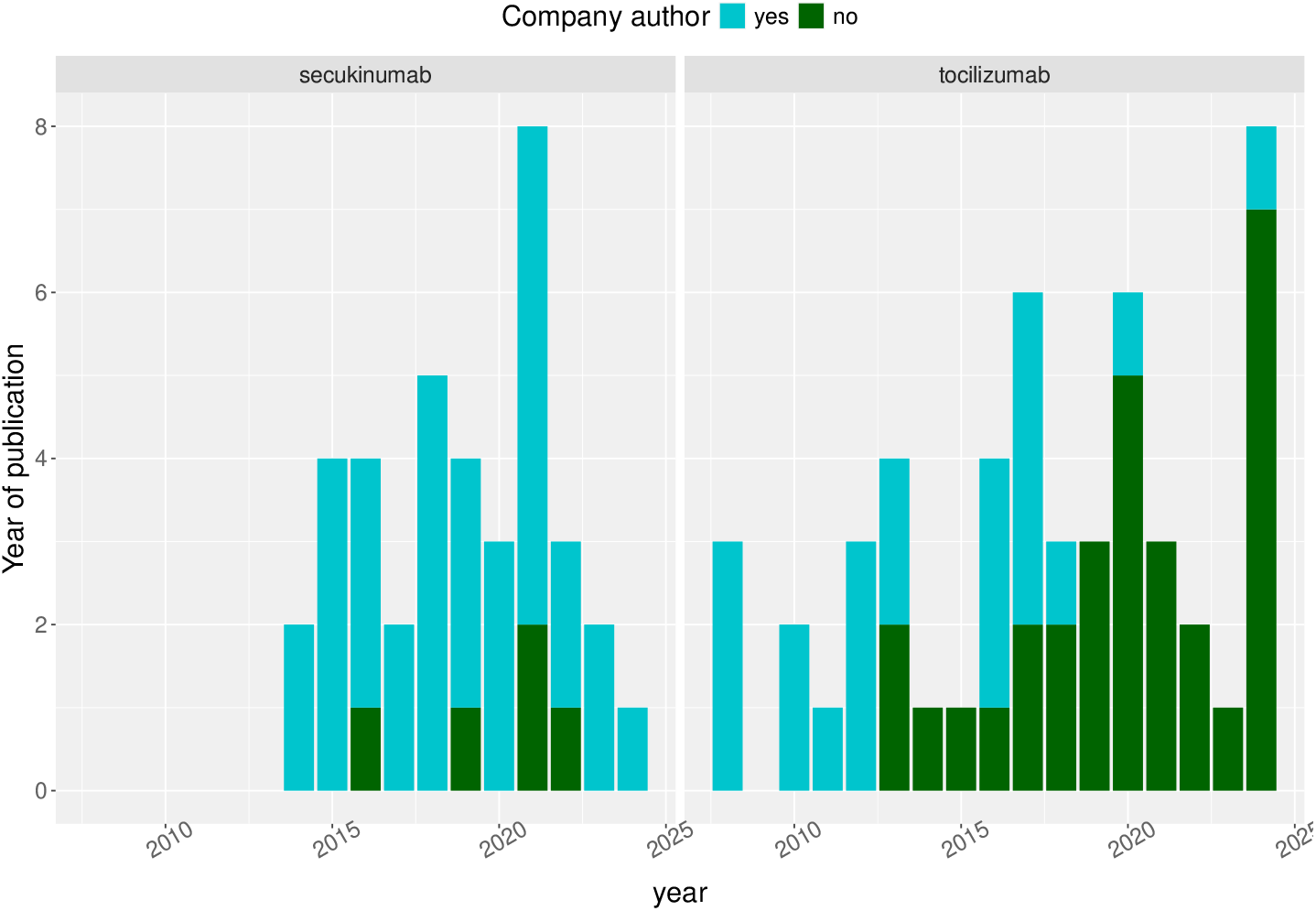
Publication year for articles studied.

Figure 3 shows the pathology considered for inclusion, and Figure 4 regroups the journals where the articles were published using a treemap. Tocilizumab was mainly tested for rheumatoid arthritis (N=44, including one study where patients were recruited from multiple indications), with one or two papers each reporting studies in other pathologies (psoriatic arthritis, systemic sclerosis, giant cell arteritis). Secukinumab was mostly used to treat patients with psoriatic arthritis (N=23, plus one paper in juvenile psoriatic arthritis), and a second indication was psiorasis (N=6) and plaque psoriasis (N=10). All the articles published in Dermatology journals were about secukinumab, because of its main indication in psoriasis. Supplementary table 3 lists the journals and which category they were ascribed to.

**Figure 3:**
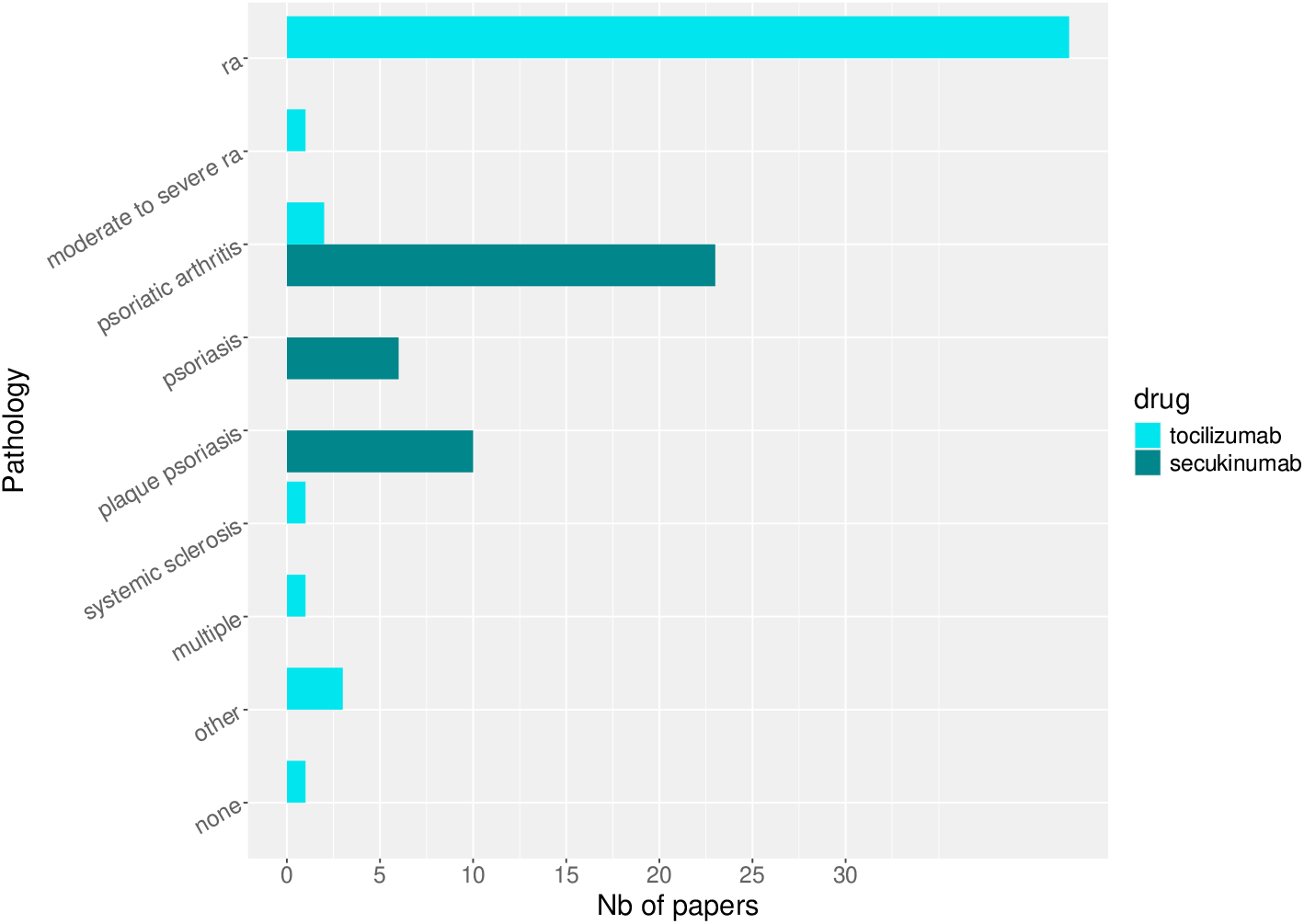
Publication type: main pathology studied in the paper.

**Figure 4:**
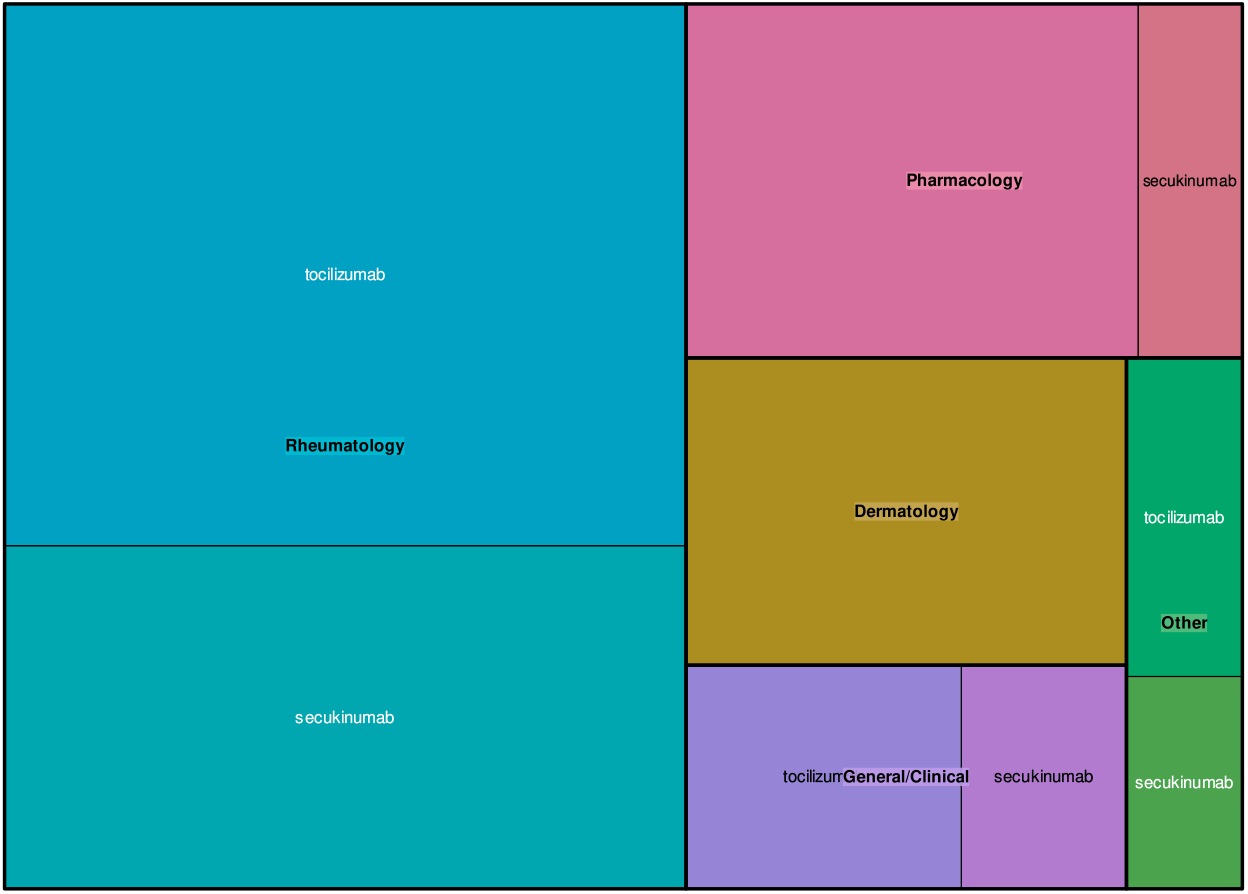
Type of journal where the paper was published, one of Rheumatology (eg: Annals of Rheumatological Diseases, Arthritis Care Research,…), Dermatology (British Journal of Dermatology, …), Pharmacology (Therapeutic Drug Monitoring, Journal of Clinical Pharmacology, …), General clinical journal (eg: Lancet, New England Journal of Medicine,…), other (Pharmacogenomics, Paediatric Drugs,…). A full list of the papers and their attribution is given in Supplementary table 3. In this graph, each group of journals is represented by a colour block, with different shades for the two drugs (only one shade is shown for papers published in Dermatology journals as only papers for secukinumab were published in those journals), and the area is proportional to the number of papers published in that group.

### 3.2 Type of trial

Figure 5 illustrates that most of the studies were performed in adults. For secukinumab, 4 studies concerned paediatric patients and one included both adult and children data. No paediatric study was found in the search for tocilizumab, although the paediatric development plan proposed a development strategy in this population. Most of the studies reported phase 3 randomised trials (35 for secukinumab, 32 for tocilizumab), with two studies for secukinumab combining data from phase 1 to 3 and one study for each drug including phase 2 and phase 3 data. For tocilizumab, one study reported the use of real-world data combined with phase 3 data. For tocilizumab, which was marketed some years before, 10 studies were performed post-marketing/phase 4. In 4 other studies, the information was not clearly reported but the studies did not seem to be RCT. Only two papers in the secukinumab list reported dose-escalation, while a few papers mentioned rescue therapy or dose modifications could occur during the trial.

**Figure 5:**
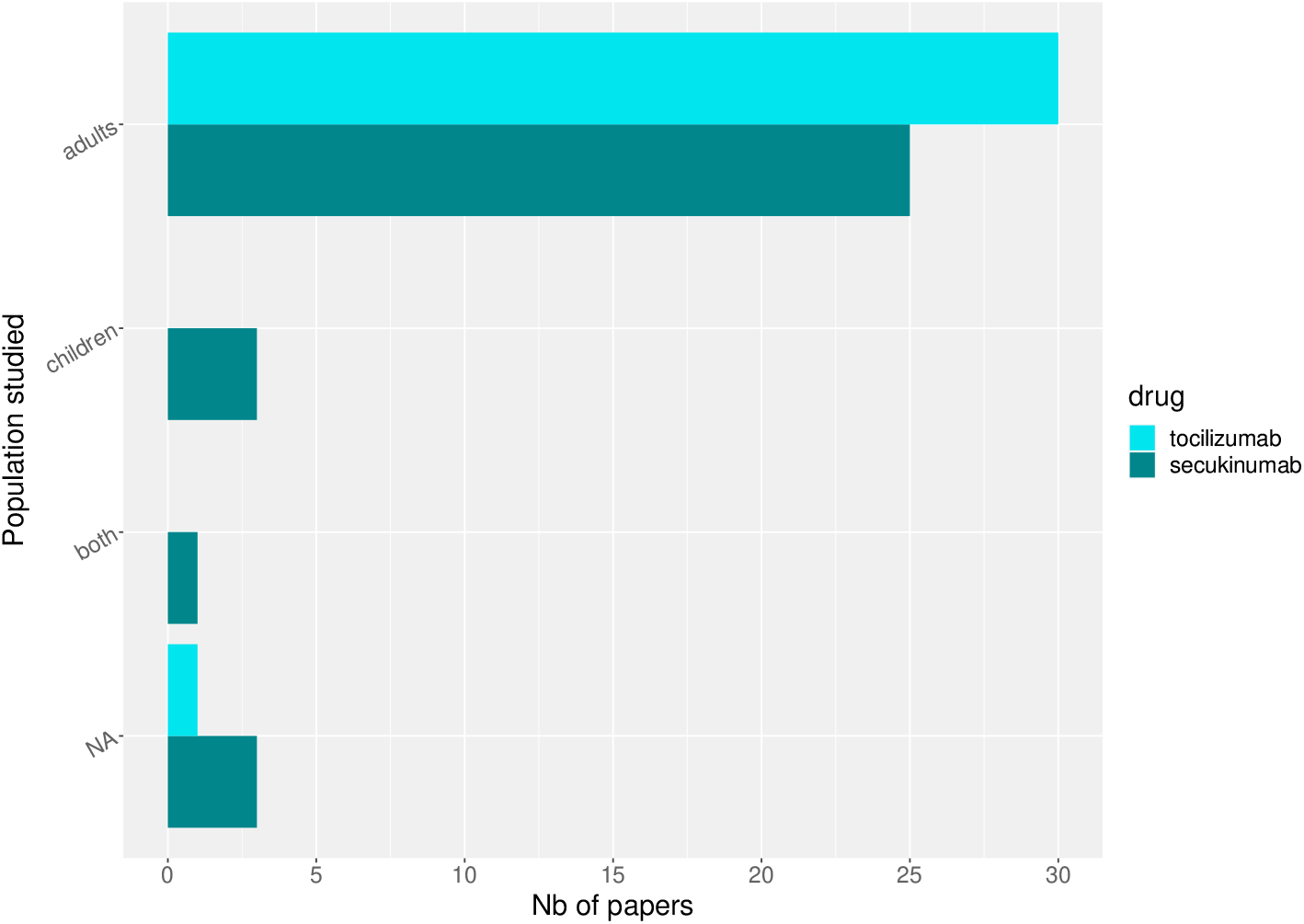
Population studied

28 papers for tocilizumab and 5 papers for secukinumab reported that the corresponding drug was given along with DMARD, but this information was missing in respectively 15 and 33 papers, and not applicable in 6 papers for tocilizumab which combined studies with monotherapy and studies where the drug was administered in combination.

#### Data analysis

Figure 6 records the main analysis performed in the study. The majority of the papers reported standard statistical analyses such as change from baseline in the primary outcome. Modelling was reported in a little over a third of the papers, but PK or PD modelling only represented a small fraction of the analyses reported.

**Figure 6:**
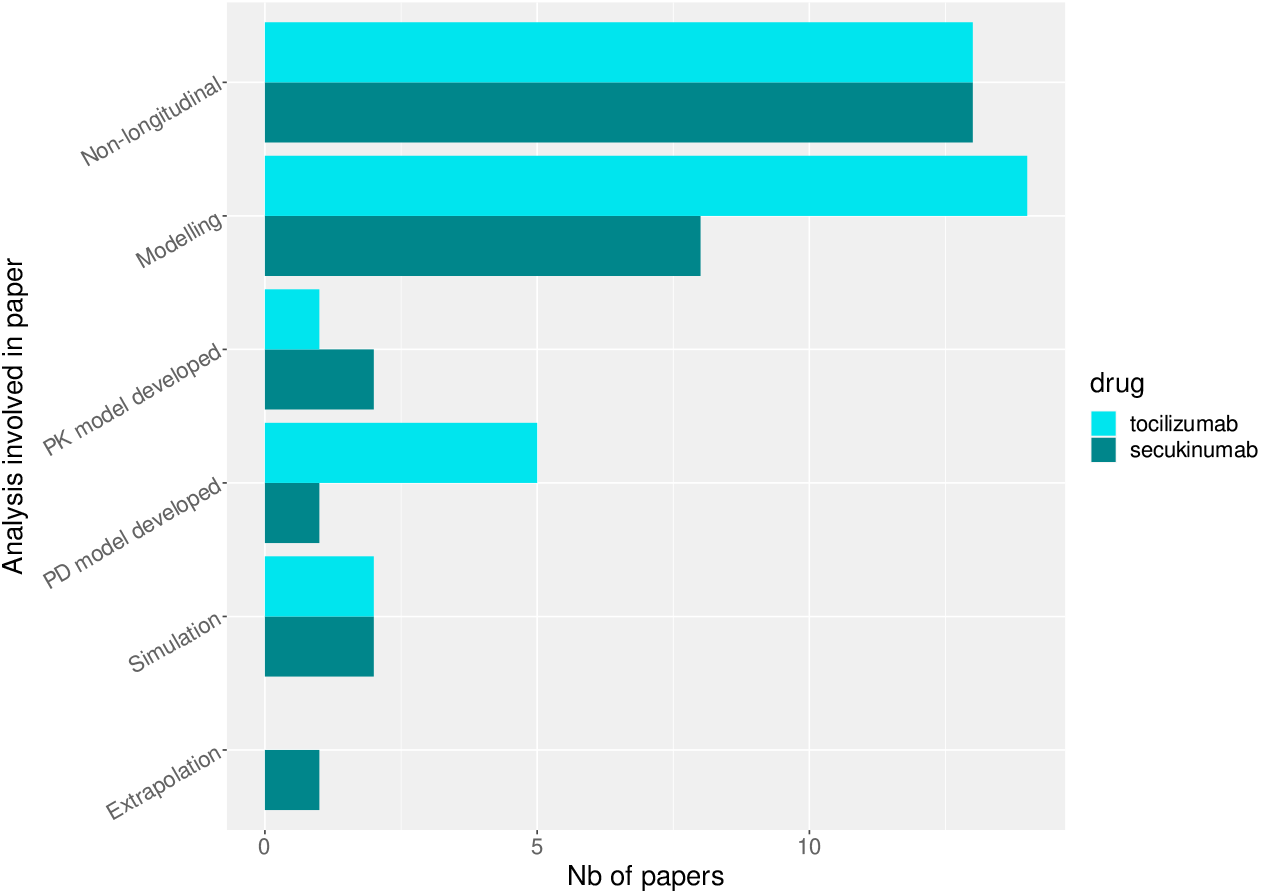
Main analysis performed in the studies.

Figure 7 summarises the main clinical questions addressed by the study. Most of the articles assessed the safety and/or efficacy of the drugs. There were few pharmacokinetic (2 for secukinumab, 6 for tocilizumab) or pharmacodynamic (4 for secukinumab, 7 for tocilizumab) studies. In addition, three papers for secukinumab used mixed models repeated measures to analyse change in repeatedly measured outcomes as a function of biomarkers and visits, but exposure was not a factor considered in these analyses. Studies investigating biomarkers or patient-reported outcomes generally considered them as change from baseline, without a longitudinal component, or evaluated the prognostic value of a biomarker (relationship between value at baseline and outcome).

**Figure 7:**
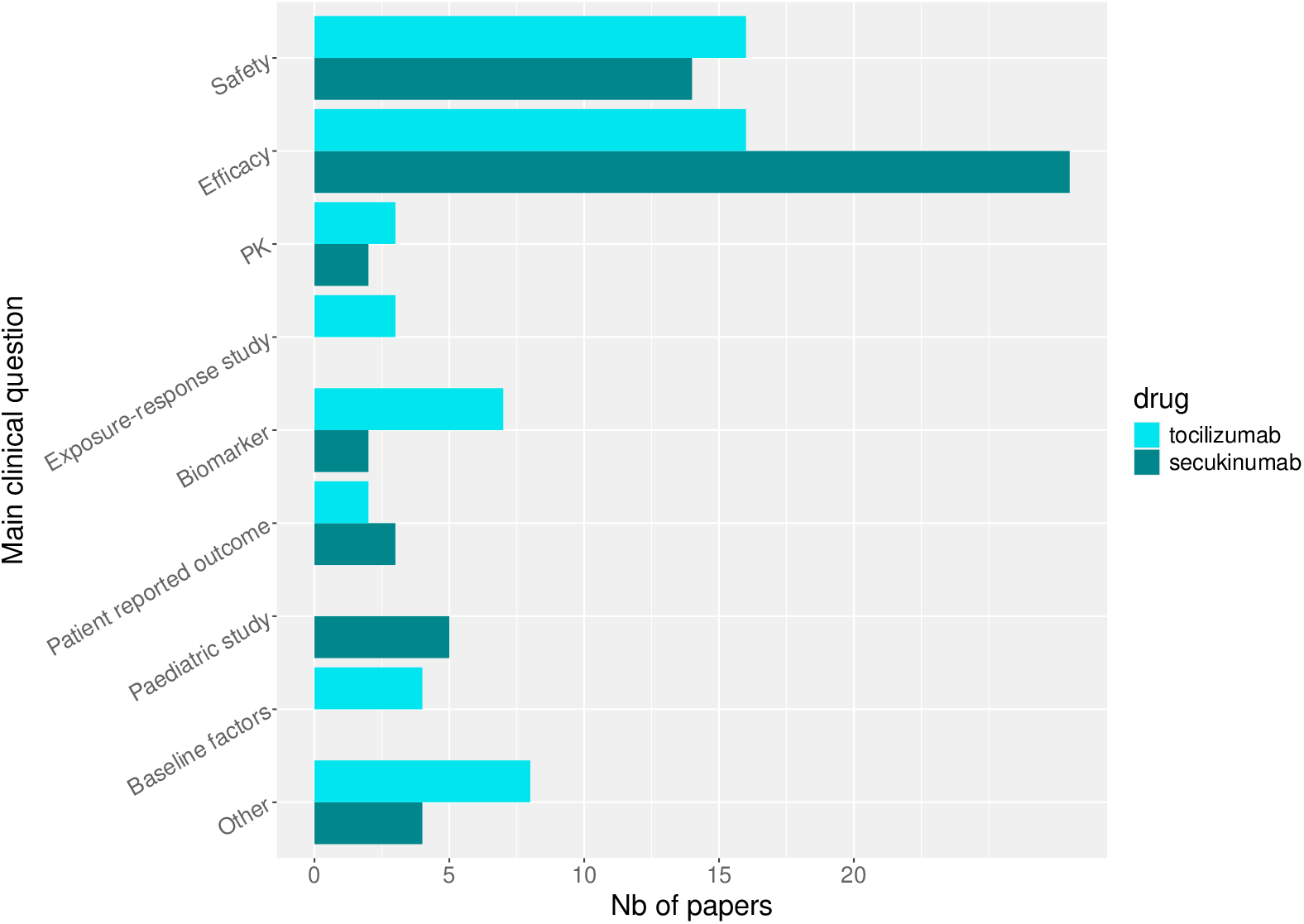
Main clinical questions (multiple choices possible).

Figure 8 lists the main outcomes used in the different studies, which included categorical outcomes (ACR, usually treated as responders to ACR20), continuous biomarkers (CRP), as well as composite scores (DAS28, HAQ, pain measured using a visual analog scale) often treated as continuous.

**Figure 8:**
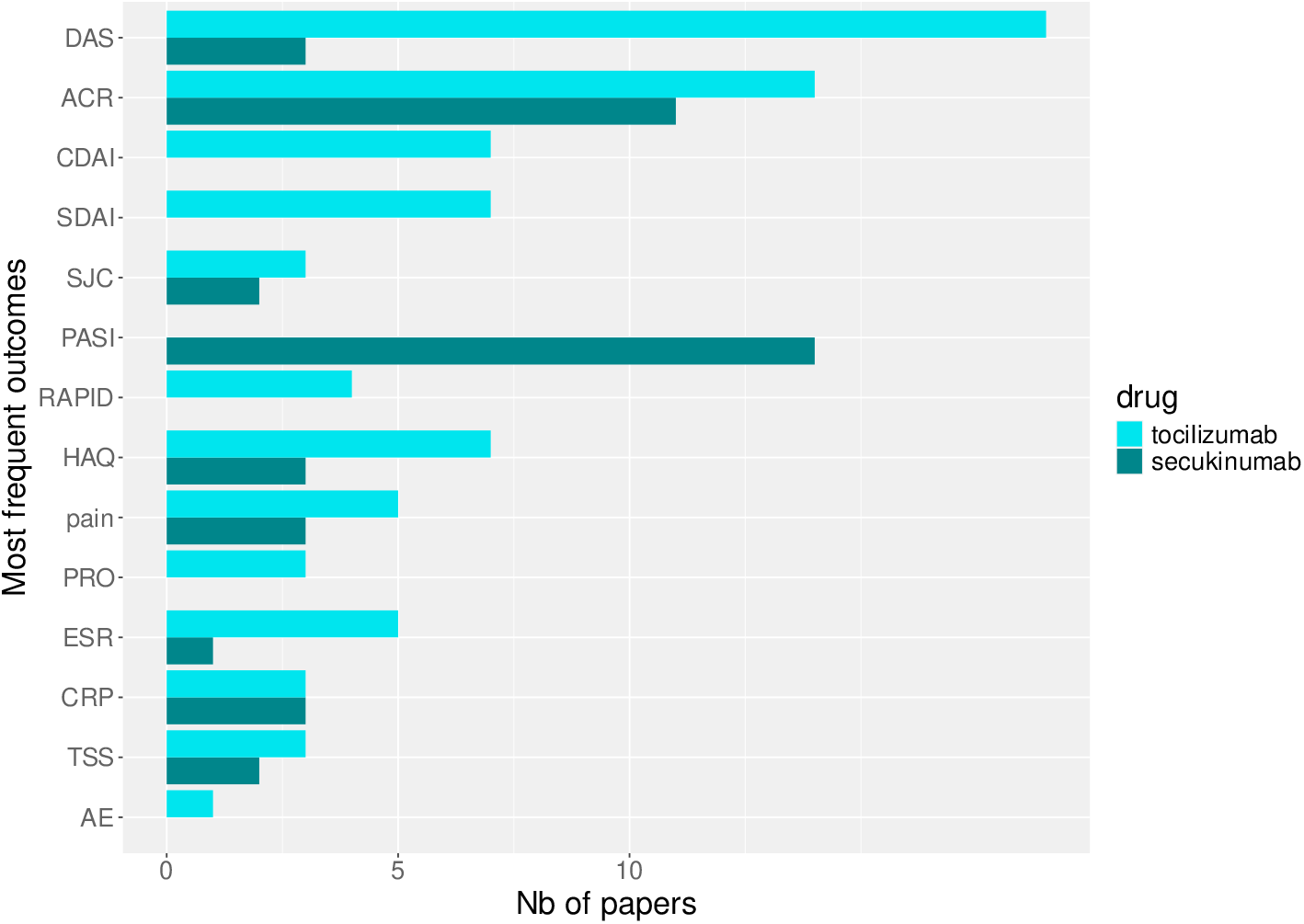
Outcomes most frequently studied (multiple choices possible).

For both drugs, many of the studies share the same base data: some use all of it, some pool several studies, some consider only specific groups, for instance the large population PK studies include patients from the phase III RCT with PK data to build a PK model. As a result, we decided not to give a number of subject in each study because some subjects are included in several datasets, and sometimes different analyses within the same paper use different sets of subjects.

Figure 9 lists the main baseline covariates mentioned in the different studies, regrouped in broad categories. Age and other demographic factors were the most often included, closely followed by disease characteristics (duration, severity), baseline values of outcome (HAQ, SJC, TJC, PASI, HQL,…), baseline biomarkers (CRP, platelet counts, ESR, C-reactive protein, IL6), comedications (methotrexate (MTX), cDMARD, previous therapies, glucocorticoids,…), comorbidities (diabetes, cardiovascular disease, renal function,…) and body size (weight, height, BMI and BSA).

**Figure 9:**
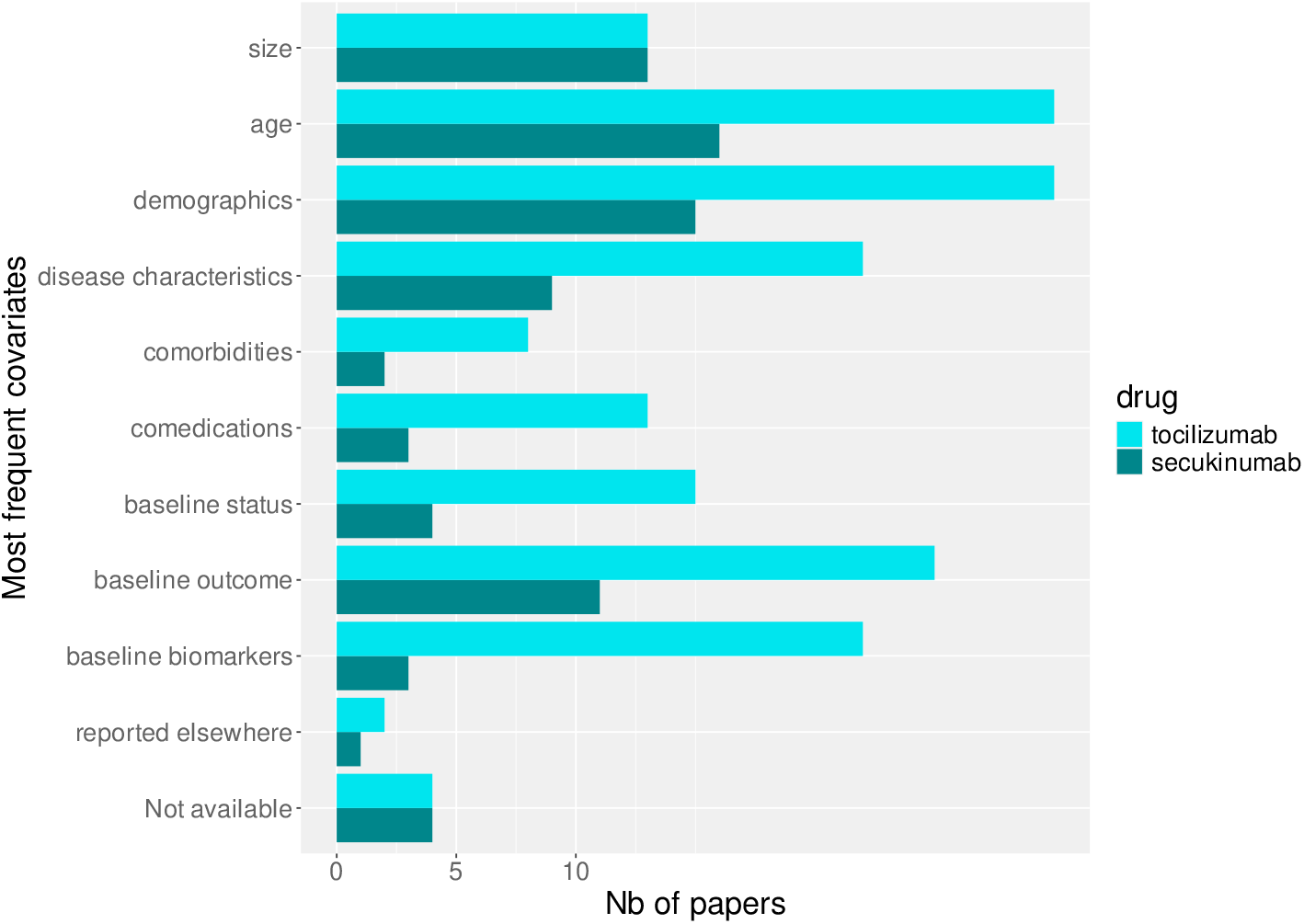
Baseline covariates most frequently mentioned (multiple choices possible).

### 3.3 PK, PD, AND PKPD MODELLING

#### PK models for secukinumab

For secukinumab, a two-compartment population PK model was developed in (Bruin et al., 2017). It was then used to simulate accumulation at steady-state with repeated dosing. A minimal PBPK model was also developed later for patients in psoriasis and coupled with compartments for binding of IL-7 in serum and skin to predict target engagement of IL-7 (Ayyar et al., 2022), The data used to develop this model was collected in 1 phase I and 3 phase II studies, as well as the 4 main phase III RCTs (ERASURE, FIXTURE, FEATURE, JUNCTURE), totalling 2730 patients. However, only mean values recovered by digitising published data and the analysis was performed by naive pooled approach Model-Based Meta-Analysis (non-linear regression of the trial level data).

#### PK models for tocilizumab

For tocilizumab, the first PK model was developed by Frey et al. (Frey et al., 2010) from nearly 1800 subjects pooled from the four main phase III RCT (AMBITION, OPTION, RADIATE, TOWARD). It included 2 compartments and the elimination was modelled with parallel linear and Michaelis– Menten processes. The model was used as is in Gibiansky and Frey to predict individual parameters (Gibiansky and Frey, 2012) and tocilizumab concentrations, entered as the exposure driver to model the pharmacodynamics in a sequential approach. The same model was then extended to an SC formulation using a first-order absorption model by (Abdallah et al., 2017).

In 2018, Bastida et al. developed a simpler PK model (Bastida et al., 2018) using a small group of 35 Spanish patients. The model included only one compartment disposition model with parallel first order (linear) and Michaelis–Menten (nonlinear) elimination kinetics. They then published a PD model (Bastida et al., 2019) using the same patients, and applied the models to evaluate dose-tapering strategies (Bastida et al., 2020).

Finally, Hooijberg et al. (Hooijberg et al., 2025) included 12 patients in a study to assess the feasibility of dose-tapering. They also derived a 2 compartment PK model with parallel linear and Michaelis–Menten elimination, using repeated trough and peak samples. The reason they give for not using the models previously described in the literature is that the patients they treat receive lower doses than previously used and the literature models do not perform adequately at low concentrations.

For completion, one of the papers read but excluded from further review was a study by Nakada and Mager which developed an integrated TMDD-CRP model to describe inflammatory responses (Nakada and Mager, 2022). The study included tocilizumab among several other molecules to propose a minimal physiologically based pharmacokinetic model for monoclonal antibodies, but the design was unclear and the relevance to tocilizumab as well.

Table 1 shows the parameter estimates reported in the 3 pop PK studies. Frey et al. (Frey et al., 2010) performed an extensive covariate search, and retained the following relationships in the final model: sex, body surface area (BSA), high-density lipoprotein cholesterol (HDL-C), and the logarithm of the rheumatoid factor were found to have an influence on CL; total protein and albumin respectively decreased and increased the volume of distribution of the central compartment, V_1_; albumin, creatinine CL, and smoking had an impact on V_*M*_. Bastida et al. found body weight (instead of BSA) and CRP to influence CL, but with large RSE in the estimation of the coefficients (Bastida et al., 2018). Finally, Hooijberg et al. included gender as a covariate for CL, fixing its value to the 20% increase in men estimated by (Frey et al., 2010), while lean body mass was found to influence both CL and V_1_ (Hooijberg et al., 2025).

**Table 1:** Comparison of the estimated population parameters for the models by Frey et al. (Frey et al., 2010), Bastida et al. (Bastida et al., 2018) and Hooijberg et al.(Hooijberg et al., 2025).

|  | Frey et al. 2010<br>(N=1800) | Bastida et al. 2018<br>(N=35) | Hooijberg et al. 2025<br>(N=12) |
| --- | --- | --- | --- |
|  | Value (%RSE) | Value (%RSE) | Value (%RSE) |
| Fixed effects |  |  |  |
| CL (L/d) | 0.3 (4) | 0.25<sup>1</sup> (24) | 0.2 (5) |
| V<sub>1</sub> (L) | 3.5 (2) | 4.83 (12) | 3.0 (5) |
| Q (L/d) | 0.2 (8) | - | - |
| V<sub>2</sub> (L) | 2.9 (5) | - | - |
| V<sub>M</sub> (mg/d) | 7.5 (6) | 5.74<sup>2</sup> (46) | 5.2 (9) |
| K<sub>M</sub> (μg/mL) | 2.7 (10) | 4.22 (79) | 0.2 (52) |
| Interindividual variability as CV% (%RSE) |  |  |  |
| CL | 39 (15) | 17 (63) | 12 (24) |
| V<sub>1</sub> | 37 (14) | 31 (26) | - |
| V<sub>2</sub> | 66 (21) | - | - |
| V<sub>M</sub> | 54 (20) | - | - |
| Residual error |  |  |  |
| Additive (μg/mL) | 2.4 (12) | 0.16 (103) | 0.1 (-) |
| Proportional (%) | 22.0 (6) | 25.5 (17) | 23.1 (7) |

CL: clearance; Q: intercompartmental clearance; V<sub>1</sub>: central volume of distribution; V<sub>2</sub>: peripheral volume of distribution; V<sub>M</sub>: maximum elimination rate. K<sub>M</sub>: Michaelis-Menten constant
<sup>1</sup> converted from L/h; <sup>2</sup> converted from mg/h. In Frey et al. (Frey et al., 2010), RSE on variances were obtained by bootstrap. That model also included correlations between CL and V<sub>1</sub>, V<sub>2</sub> and V<sub>M</sub> ranging from -0.1 to 0.6 (see publication for details) while the other two models didn't estimate correlation terms.

Other published work did not include PK drivers. Empirical models of DAS28 as a function of time were used in Leil et al.: they applied a meta-analytical approach to characterise the change in DAS28 across time, using non-linear mixed-effects modeling approach to take into account inter-trial variability in data pooled from 130 randomized, controlled clinical trials (197 trial arms) of seven approved RA drugs with different modes of action (Leil et al., 2021). The data used in this study consisted in longitudinal summary-level data records, representing data from 27,355 patients. Blair et al. used a latent class mixture modelling approach to identify different response trajectories in 485 RA patients receiving tocilizumab in the LITHE study (Blair et al., 2020), with DAS28 as the main outcome. They also applied linear mixed effects modelling to study the change in serological biomarkers from baseline but did not detect any baseline demographic covariate or predictive biomarkers for the 3 response groups identified, only differences in baseline DAS28, HAQ and tender joint counts.

#### PD and PK/PD models for secukinumab

None of the papers identified in our review actually included a non-linear mixed effect PK/PD model for secukinumab. The only paper relating PK to an outcome is the PD model for IL-7 in serum and skin was included in the minimal PBPK model developed by Ayyar et al. (Ayyar et al., 2022).

A paper by Falck et al proposed a methodology for co-analysis of multiple endpoints using a latent variable approach, applying it to secukinumab in RA and psoriasis (Falck et al., 2024). The endpoints considered included the number of tender joints, the number of swollen joints, Physian VAS, patient VAS, SF36-MCS, SF36-PCS, HAQDI, DAS28ERS, DAS28CRP, ERS, CRP, and pain. However, PK was not part of the factors considered.

#### PD and PK/PD models for tocilizumab

Gibiansky and Frey used the PK model developed in (Frey et al., 2010) to develop a PK/PD model for sIL-6R as a function of tocilizumab concentrations (Gibiansky and Frey, 2012). They then investigated the relationship between unbound sIL-6R and neutrophil (direct effect) or platelet counts (transit-comp life span model) to explore the exposure-safety relationship. Data from the TOWARD and OPTION trials (N=2243 patients) was also used to develop an indirect response model with sigmoid inhibition of DAS28 production as a function of the AUC of tocilizumab by Levi, Grange and Frey (Levi et al., 2013). Eight covariates were included in the final model, with the following relationships: an inverse relationship between EC50 and log-transformed IL-6; higher Emax in males; higher Kout in blacks and whites combined compared with other races; positive relationships between BASE and Health Assessment Questionnaire (HAQ), log-transformed IL-6, PAIN, and physician’s global score of disease activity (VASP); and an inverse relationship between background DMARD effect in the control group and log-transformed IL-6. Bastida et al. proposed a PD model for DAS28 as a function of concentrations (Bastida et al., 2019) instead of AUC, also modelling SDAI and CDAI. They developed a latent variable model applied to biomarkers of inflammation and disease scores.

### 3.4 Summary of the quantitative review

Coming back to the research questions we set out to address in this study, we find the following key messages. The clinical questions of interest were mostly centered on efficacy and safety. As a result, most of the primary analyses were standard statistical analyses (endpoint comparison, target achievement), with very classical estimands such as the comparison of a clinical score after one year of treatment. For both drugs, many of the studies share the same base data variables. There were very few longitudinal data analyses performed, and even in those papers information on study design concerning the number and timing of samples was hard to come by, in part because most of these studies pooled data from several RCT with different designs. Several papers published PK models in very small studies with more detailed descriptions of the design, and did not use information about models developed in much larger populations (in particular, no use of such models as priors).

Most papers we found included phase III patients, with some studies combining data from phase I to III, and most of the modelling shared the same datasets (with the exception of a couple very small PK and PK/PD studies in the case of tocilizumab, which were performed in monocentric settings). A couple of papers used real-world data but only as follow-up of patients included in registry and cohorts, and we excluded those which involved comparison between treatment strategies with different drugs as they did not specifically consider tocilizumab or secukinumab. Ghuman et al. used real-world data from the Royal Free Scleroderma Cohort(SMART), a prospective observational cohort of SSc patients who consented to research, to study the potential risk factors for lung function decline in patients with SSc-ILD(Ghuman et al., 2024).

We found very few examples of extrapolation approaches studying differences in patient characteristics or disease characteristics. One paper in the secukinumab group presented a methodology to extrapolate across disease severity and age groups, but we found no example of papers extrapolating across differences in ethnicity, disease types (such as juvenile versus rheumatoid arthritis), or disease characteristics such as different progression rates.

When available, the types of longitudinal models developed to describe the data were mostly PK and PK/PD, with a couple of clustering approaches such as latent class modelling. PK models were very classical, and estimation was almost always performed with the NONMEM software. PD models were also quite standard for tocilizumab, with mostly direct or indirect response models for DAS28. They were a bit more creative for secukinumab, with no actual PK/PD model developed in the papers we found, but one example of a minimal PBPK model including compartments for an efficacy biomarker, and one model using a latent variable approach to analyse multiple endpoints. We found very few examples of extrapolation, and notably only one paper extrapolating across disease and age groups. Simulation studies were almost always either used to produce diagnostic graphs for model evaluation, or to simulate different dosing regimen. A series of simulation studies for tocilizumab evaluated dose-tapering strategies for long-term patients with the aim to reduce the drug burden.

### Limits of this work

initially the list was provided by the case-study owners and based on phase 3 data so a limited number of publications dealt with PK and PD. Although further searches expanded to specifically target PK and PD models, we didn’t manage to recover many papers on modelling, and very few used longitudinal data for extrapolation or advanced simulations. More general keyword searches led to large numbers of hits which were intractable within WP1.1. A suggestion was made to use AI for selection but this could not be implemented as there was no manpower for this, and we would still need to sift manually through at least titles and usually abstracts.

In particular none of the papers for tocilizumab included in the first search, centered around the 4 large RCT in RA patients, had PK models in them, and the first PK model was only found using the list of references from initially included papers. This prompted the second and third keyword searches which were specifically targeted at PK and PK/PD models, but even with those we found about 10 papers for tocilizumab and 5 papers for secukinumab relevant to our work.

Concerning regulatory documents, the registration file included an enormous amount of information but not much additional published material relevant to the purposes of modelling and simulation. We lacked the resources to exhaustively track all the references from the registration file but this could be a perspective of this work. In terms of paediatric studies, we were disappointed to find very little published material. The PIP included very limited information listing the steps to be undertaken and were more of a roadmap. Some studies were later performed, some weren’t. When they were published we included them in the analysis.

## 4 Discussion & Conclusions

The objective of this work was to review the modelling and simulation methods used in the two selected case-studies, with a focus on longitudinal data modelling, simulation methods and extrapolation approaches (across populations, across indications or across similar drugs) applied during the later phases of drug development. Unfortunately, despite the clear focus on longitudinal modelling and simulation in our search, we found very few examples of PK or PK/PD models in our search: even when outcomes were measured repeatedly during the study, the focus of the analyses was usually along the lines of improvement at a given time-point, with other time-points included in secondary analyses. When PK or PK/PD models were developed, they were generally simple, with compartmental models for PK and direct or indirect response models for PD.

For tocilizumab, the first PK model published included just about all the patients from phase III studies, and this amount of data could only be obtained by Roche authors. As expected, we saw a trend towards most of the early publications (in terms of year of development) involving company co-authors as data is first collected in-house, while later publications were more often performed outside the company developing the drug. This could also be the reason why not much novel modelling was found in our review, as it is possible that that work would have been done in house and not published, or not published yet. A similar reason could also explain why we found almost no paediatric study, despite modelling and simulation analyses being put forward in the Paediatric Investigation Plan. Another reason could be that the planned studies are not completed yet, or that waivers have been obtained and the studies were never done. Finally, sensitive modelling could also still remain unpublished.

Clinical questions addressed in the studies selected in this review were mainly centered on efficacy (57%, including exposure-response relationships and efficacy concerning patient-reported outcomes) and safety (34%). Besides the few studies investigating PK, other clinical questions focused on finding predictors of response, either biomarkers related to changes in the outcome under study, molecular characteristics of certain groups of patients, or prognostic factors at baseline. Finally, a couple of studies were more methodological, including an extrapolation across both disease severity and age-groups, as well as a study analysing multiple endpoints through latent variables.

## Data Availability

All data produced in the present study are available upon reasonable request to the authors.

## Acknowledgments

This work was performed within the project INVENTS (https://invents-he.eu/), which has received funding from the European Union’s Horizon Europe Research and Innovation programme under grant agreement 101136365, the Swiss State Secretariat for Education, Research and Innovation (SERI) and by the UKRI Innovative UK under their Horizon Europe Guarantee scheme.

## Supplementary material

### Keyword searches

Table 2 gives the keyword search performed to expand the list of papers. For both molecules, the first search gave too many hits, most of which appeared irrelevant from scanning the first abstracts in the list. The other searches had most articles in common, and were combined to expand the initial list of papers, reading the abstract to determine whether they were relevant and should be included.

**Table 2:** Expanded keyword searches, focusing on modelling, simulation, extrapolation, PK and PD analyses. Searches 1 to 6 for tocilizumab, and 1 to 8 for secukinumab, were combined to form the full list of articles considered for the review.

| Search | Keyword search | Nb hits |
| --- | --- | --- |
| **Tocilizumab** |  |  |
| 0 | tocilizumab AND (modelling OR modeling OR extrapolation OR simulation OR longitudinal OR repeated OR NONMEM OR Monolix OR "non-linear mixed effect" OR pharmacokinetic* OR pharmacodynamic*) AND ("rheumatoid arthritis" OR "systemic sclerosis") | 398 |
| 1 | tocilizumab AND (longitudinal) AND (modelling OR modeling) | 28 |
| 2 | tocilizumab AND (modelling OR extrapolation OR simulation) AND (longitudinal OR repeated OR NONMEM OR Monolix OR "non-linear mixed effect") AND (rheumatoid arthritis) | 24 |
| 3 | tocilizumab AND (modelling OR modeling OR extrapolation OR simulation OR longitudinal OR repeated OR NONMEM OR Monolix OR "non-linear mixed effect" OR pharmacokinetic* OR pharmacodynamic*) AND (systemic sclerosis) | 17 |
| 4 | tocilizumab AND (pharmacokinetic* OR pharmacodynamic* OR toxicity OR efficacy OR biomarker) AND (longitudinal OR repeated OR NONMEM OR Monolix OR "non-linear mixed effect") AND ("rheumatoid arthritis" OR "systemic sclerosis") | 18 |
| 5 | tocilizumab AND (rheumatoid arthritis) AND pharmacokinetic* AND model | 25 |
| 6 | tocilizumab AND (rheumatoid arthritis) AND (pharmacodynamic*) AND model | 14 |
| **Secukinumab** |  |  |
| 1 | secukinumab AND (longitudinal) AND (modelling OR modeling) | 8 |
| 2 | secukinumab AND (modelling OR extrapolation OR simulation) AND (longitudinal OR repeated OR NONMEM OR Monolix OR "non-linear mixed effect") AND ("psoriasis" OR "Systemic juvenile idiopathic arthritis" OR "enthesitis-related arthritis") | 6 |
| 3 | secukinumab AND (modelling OR modeling OR extrapolation OR simulation) AND (longitudinal OR repeated OR NONMEM OR Monolix OR "non-linear mixed effect") AND ("psoriasis" OR "Systemic juvenile idiopathic arthritis" OR "enthesitis-related arthritis") | 6 |
| 4 | secukinumab AND (pharmacokinetic* OR pharmacodynamic* OR toxicity OR efficacy OR biomarker) AND (longitudinal OR repeated OR NONMEM OR Monolix OR "non-linear mixed effect") AND ("psoriasis" OR "Systemic juvenile idiopathic arthritis" OR "enthesitis-related arthritis") | 15 |
| 5 | secukinumab AND (Psoriatic Arthritis) AND pharmacokinetic* AND model | 1 |
| 6 | secukinumab AND (Psoriatic Arthritis) AND pharmacodynamic* AND model | 0 |
| 7 | secukinumab AND (Psoriasis) AND pharmacokinetic* AND model | 6 |
| 8 | secukinumab AND (Psoriasis) AND pharmacodynamic* AND model | 9 |

### Journal categories

Table 3 lists the journals and which category they were ascribed to, and shows the number of papers published in each journal for the two drugs being studied.

**Table 3:**
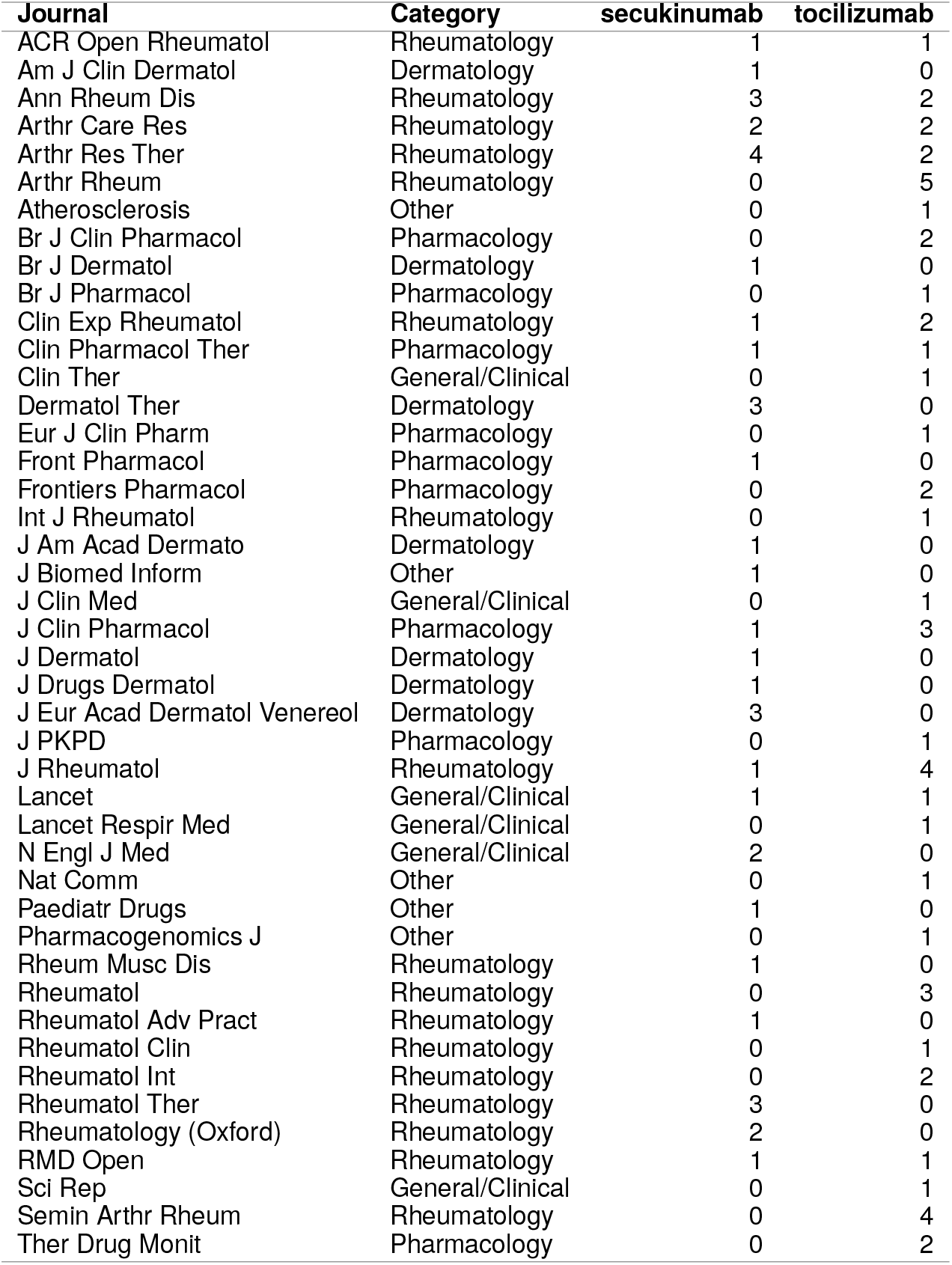
List of journals with the number of articles published in each of them for both drugs in the case-studies.

| Journal | Category | secukinumab | tocilizumab |
| --- | --- | --- | --- |
| ACR Open Rheumatol | Rheumatology | 1 | 1 |
| Am J Clin Dermatol | Dermatology | 1 | 0 |
| Ann Rheum Dis | Rheumatology | 3 | 2 |
| Arthr Care Res | Rheumatology | 2 | 2 |
| Arthr Res Ther | Rheumatology | 4 | 2 |
| Arthr Rheum | Rheumatology | 0 | 5 |
| Atherosclerosis | Other | 0 | 1 |
| Br J Clin Pharmacol | Pharmacology | 0 | 2 |
| Br J Dermatol | Dermatology | 1 | 0 |
| Br J Pharmacol | Pharmacology | 0 | 1 |
| Clin Exp Rheumatol | Rheumatology | 1 | 2 |
| Clin Pharmacol Ther | Pharmacology | 1 | 1 |
| Clin Ther | General/Clinical | 0 | 1 |
| Dermatol Ther | Dermatology | 3 | 0 |
| Eur J Clin Pharm | Pharmacology | 0 | 1 |
| Front Pharmacol | Pharmacology | 1 | 0 |
| Frontiers Pharmacol | Pharmacology | 0 | 2 |
| Int J Rheumatol | Rheumatology | 0 | 1 |
| J Am Acad Dermatol | Dermatology | 1 | 0 |
| J Biomed Inform | Other | 1 | 0 |
| J Clin Med | General/Clinical | 0 | 1 |
| J Clin Pharmacol | Pharmacology | 1 | 3 |
| J Dermatol | Dermatology | 1 | 0 |
| J Drugs Dermatol | Dermatology | 1 | 0 |
| J Eur Acad Dermatol Venereol | Dermatology | 3 | 0 |
| J PKPD | Pharmacology | 0 | 1 |
| J Rheumatol | Rheumatology | 1 | 4 |
| Lancet | General/Clinical | 1 | 1 |
| Lancet Respir Med | General/Clinical | 0 | 1 |
| N Engl J Med | General/Clinical | 2 | 0 |
| Nat Comm | Other | 0 | 1 |
| Paediatr Drugs | Other | 1 | 0 |
| Pharmacogenomics J | Other | 0 | 1 |
| Rheum Musc Dis | Rheumatology | 1 | 0 |
| Rheumatol | Rheumatology | 0 | 3 |
| Rheumatol Adv Pract | Rheumatology | 1 | 0 |
| Rheumatol Clin | Rheumatology | 0 | 1 |
| Rheumatol Int | Rheumatology | 0 | 2 |
| Rheumatol Ther | Rheumatology | 3 | 0 |
| Rheumatology (Oxford) | Rheumatology | 2 | 0 |
| RMD Open | Rheumatology | 1 | 1 |
| Sci Rep | General/Clinical | 0 | 1 |
| Semin Arthr Rheum | Rheumatology | 0 | 4 |
| Ther Drug Monit | Pharmacology | 0 | 2 |

